# Treatment practice of vasospasm during endovascular thrombectomy: an international survey

**DOI:** 10.1101/2023.08.01.23293498

**Authors:** J. Jesser, T. N. Nguyen, A. A. Dmytriw, H. Yamagami, Z. Miao, L. J. Sommer, A. Stockero, J. A. R. Pfaff, J. M. Ospel, M. Goyal, A. B. Patel, V. Mendes Pereira, U. Hanning, L. Meyer, W. van Zwam, M. Bendszus, M. Wiesmann, M. A. Möhlenbruch, C. S. Weyland

**Author notes:** Corresponding author: PD Dr. Charlotte S. Weyland Department of Neuroradiology, University Hospital RWTH Aachen Pauwelsstraße 30 52076 Aachen Germany.

## Abstract

**Background and aim:** The clinical importance and management of vasospasm as a complication during endovascular stroke treatment (EVT) has not been well studied. We sought to investigate current expert opinions in neuro-intervention and therapeutic strategies of iatrogenic vasospasm during EVT.

**Methods:** We conducted an anonymous international online survey (April 04^th^ to May 15^th^ 2023) addressing treatment standards of neurointerventionalists (NI) practicing EVT. Several illustrative cases of patients with vasospasm during EVT were shown. Two study groups were compared according to the NI’s opinion regarding the potential influence of vasospasm on patient outcome after EVT using descriptive analysis.

**Results:** In total, 534 NI from 56 countries responded, of whom 51.5% had performed more than 200 EVT. Vasospasm was considered a complication potentially influencing the patient’s outcome by 52.6% (Group 1) whereas 47.4% did not (Group 2). Physicians in Group 1 more often added vasodilators to their catheter flushes during EVT routinely (43.7% vs. 33.9%, p = 0.033) and more often treated severe large-vessel vasospasm with vasodilators (75.3% vs. 55.9%; p < 0.001), as well as extracranial vasospasm (61.4% vs. 36.5%, p < 0.001) and intracranial medium-vessel vasospasm (27.1 % vs. 11.2%, p < 0.001), compared to Group 2. In case of a large-vessel vasospasm and residual and amenable medium vessel occlusion during EVT, the study groups showed different treatment strategies. Group 2 continued the EVT immediately more often, without initiating therapy to treat the vasospasm first (9.6% vs 21.1%, p < 0.001).

**Conclusion:** There is disagreement among neurointerventionalists about the clinical relevance of vasospasm during EVT and its management. There was a higher likelihood of use of preventive and active vasodilator treatment in the group that perceived vasospasm as a relevant complication as well as differing interventional strategies for continuing an EVT in the presence of a large-vessel vasospasm.

## Introduction

More than 10% of endovascular stroke treatments (EVTs) for acute ischemic stroke are associated with perioperative complications [1]. They include distal embolization to a new territory (4-6%), de novo stenosis of target vessels (3.4%), vessel perforation (0.6-4.9 %), dissection (0.6 – 3.9%), groin hematoma (2 – 10%) and vasospasm (3.9 – 23%) [2–4]. Iatrogenic vasospasm is a common complication that occurs during stent retriever or contact aspiration instrumentation due to the vessel wall irritation during thrombectomy maneuvers. On angiography, vasospasm can be perceived as a concentric contraction of the arterial vessel wall.

Vasospasm is more likely to occur in younger stroke patients as well as in EVT with multiple thrombectomy attempts [6]. While vasospasm during EVT is considered as non-serious and a transient complication by some authors [7], others discuss vasospasm as a potential cause for stroke recurrence [8]. The clinical relevance of vasospasm as a complication during EVT remains uncertain as well as geographical differences in incidence rates or related clinical sequelae. A higher prevalence of coronary vasospasm is demonstrated in Asian populations compared to the Western World [9]. While racial differences in vasomotor reactivity after acute myocardial infarction are being discussed for several years [10] and genetic determinants of vasospasm after subarachnoid hemorrhage have been defined [11], regional differences have not been investigated for cerebral vasospasm during EVT yet.

Intra-arterial application of vasodilators, such as calcium channel blockers (CCBs), can resolve vasospasm in most cases [12]. CCBs can be added to catheter flushes to prevent vasospasm during EVT or they can be given intra-arterially via the intermediate or guide catheter after detection of vasospasm. Withdrawal of the catheter from the affected vessel and waiting is another strategy to manage vasospasm. As there are no guidelines available to guide management for vasospasm during EVT, we aimed to define the current treatment practice for vasospasm during EVT among neuro- interventionalists.

## Methods

This was an international, anonymous online survey conducted from April 4 until May 15, 2023. Ethics approval was granted by the local research ethics board (Medical Faculty, University of Aachen, Germany, local registration number 23-095). In total, 18 questions were developed to evaluate treatment strategies of vasospasm during EVT by neuro-interventionalists with a survey duration of approximately five minutes. Angiographic images of three patients with vasospasm in different territories (extracranial internal carotid artery, proximal middle cerebral artery M1 segment, distal medium vessel segment) were shown and respondents asked to answer whether they would treat with intra-arterial vasodilator or not. The complete survey questions are listed in the supplement. Analysis and reporting followed the recommendations of the Consensus –Based Checklist for Reporting of Survey Studies (CROSS) [13].

The survey was distributed at a neuro-interventional conference (World Live Neurovascular Conference 2023), via electronic communication of the German Society for Neuroradiology (distributed to 1450 members) and the European Society of Minimally Invasive Neurological Therapy (distributed to 3438 members, e-mail opened by 33%, link opened by 5.4%), and via invitation by co-authors to their global colleagues. Internet Protocol (IP) – addresses were anonymously saved by the survey’s online platform to prevent duplicate response bias.

## Statistical Analysis

Data are shown as number of events and percentage (n, %). After testing for normal distribution with the Shapiro-Wilk Test, further analysis was conducted with the Mann-Whitney-U-Test or χ2 test to compare groups, as appropriate. All tests were performed on the basis of a two-sided level of p-value of less than 0.05 considered statistically significant. P-values were corrected for a false discovery rate of 0.05 (Benjamini-Hochberg adjusted P-values). Statistical analyses were performed by using SPSS Statistics (29.0; IBM, Armonk, NY).

## Results

In total, 534 neuro-interventionalists (NI) from 58 countries participated in the survey with most participants (n, %) from China (109, 20.4%), Germany (66, 13.2%) and the USA (53, 10.3%) – number of participants per country available as supplemental material. The estimated response rate ranges at 20%. The majority of participants was male (77.3%, 22.4% women, 0.6% diverse). Regarding specialty, the respondents comprised of interventional neuroradiologists (50.3%), interventional neurologists (28.6%), endovascular surgeons (13.6%), interventional radiologists (3.6%) and other (3.9%). Most participants were experienced with more than 10 years of experience in endovascular stroke treatment (EVT) in 42.1% (5 – 10 years in 28.3%, 1 – 5 years in 25.1% and less than 1 year in 4.5 %) and more than 200 EVT performed in 51.4 % (100–200 EVT in 17.4%, 50–100 EVT in 14.2%, less than 50 EVT in 17.0%).

A slight majority of respondents, 52.62% (n = 281) considered vasospasm as a complication potentially influencing the patient’s outcome (Group 1) while 47.4% (n = 253) did not (study group 2). Group 2 respondents tended to be more experienced with > 200 EVTs and male respondents tended to consider vasospasm more often as relevant complication than female respondents (Group 1 – 54.6 % of male respondents and 46.6 % of female respondents) – see Table 1. Respondents currently practicing in North America considered vasospasm less frequently as relevant complication (Group 1 n (%): 26 (9.3) vs. Group 2 42 (16.6), p = 0.033) while respondents practicing in Asia tended to consider vasospasm as relevant complication (Group 1 n (%): 107 (38.1) vs. 78 (30.8), p = 0.160). Respondents from Asia added vasodilators more often routinely to their flushes regardless of their opinion on vasospasm being a relevant complication (Participants practicing in Asia: 84/186 45.2% vs. others: 126/348 36.2%, p = 0.027). Study groups differed regarding treatment strategies for vasospasm during EVT. Group 2 was more likely not to add vasodilators to their catheter flushes to prevent vasospasm compared to Group 1 (66.1% vs. 56.2%, p = 0.03). Group 2 was more likely than Group 1 to immediately continue EVT to treat an amenable medium vessel occlusion if a proximal vessel vasospasm was visualized without treating the vasospasm (21.1% vs. 9.6%, p = 0.008). More NI in Group 2 would wait for spontaneous regression of vasospasm and then reassess (21.9 % vs. 12.8 %, p < 0.001). Regarding contraindications for medical treatment of vasospasm during EVT, Group 1 considered suspected extended infarction more often as possible contraindication (60.0 % vs. 19.9 %, p = 0.007), while Group 2 more often than Group 1 did not consider any potential contraindication as relevant (19.9%, vs. 13.6 %, p = 0.034). Group 1 had more often a standard operating procedure (SOP) on treating vasospasm during EVT in place at their facility (32.1 % vs. 17.9 %, p < 0.001, Table 1).

**Table 1.**
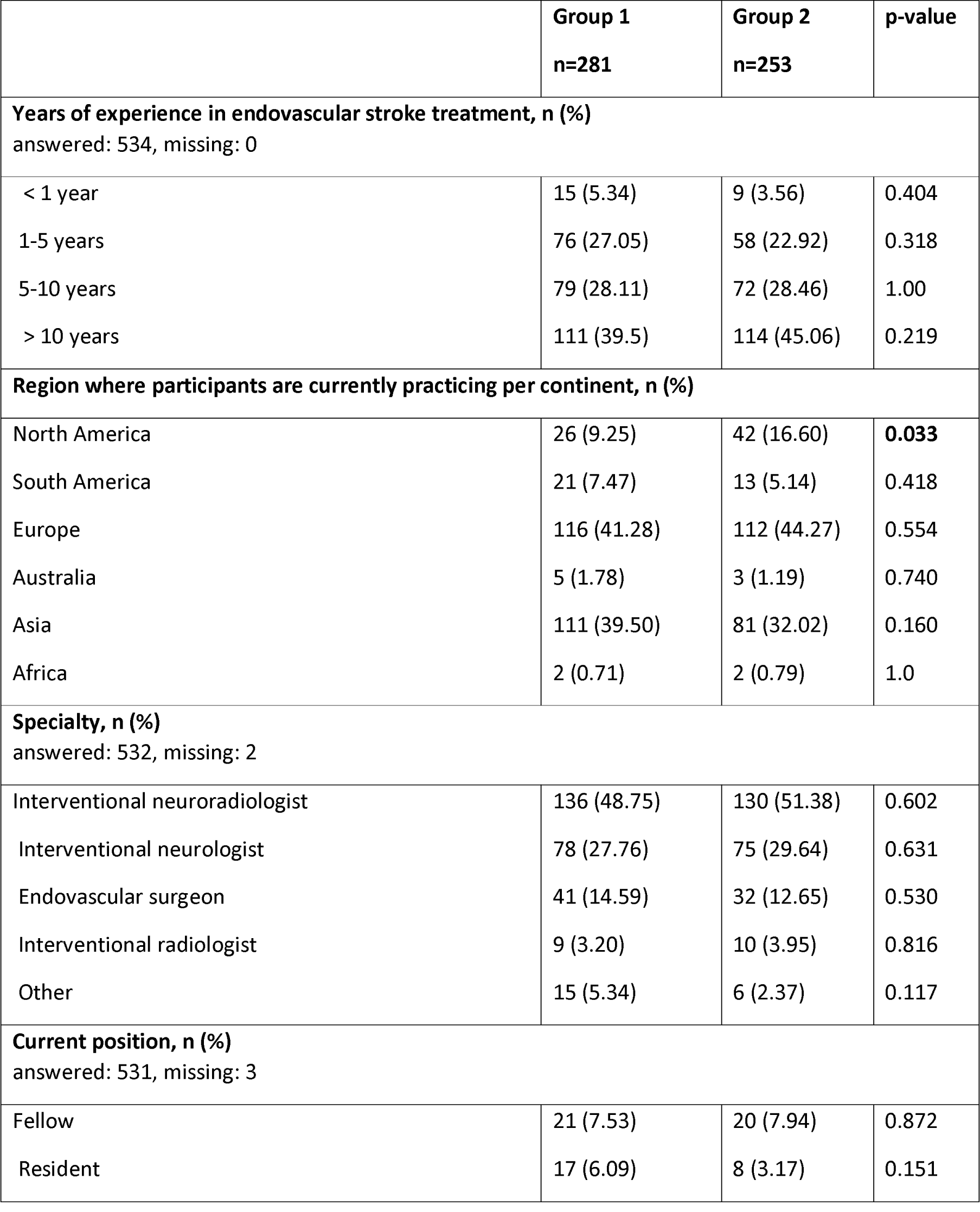
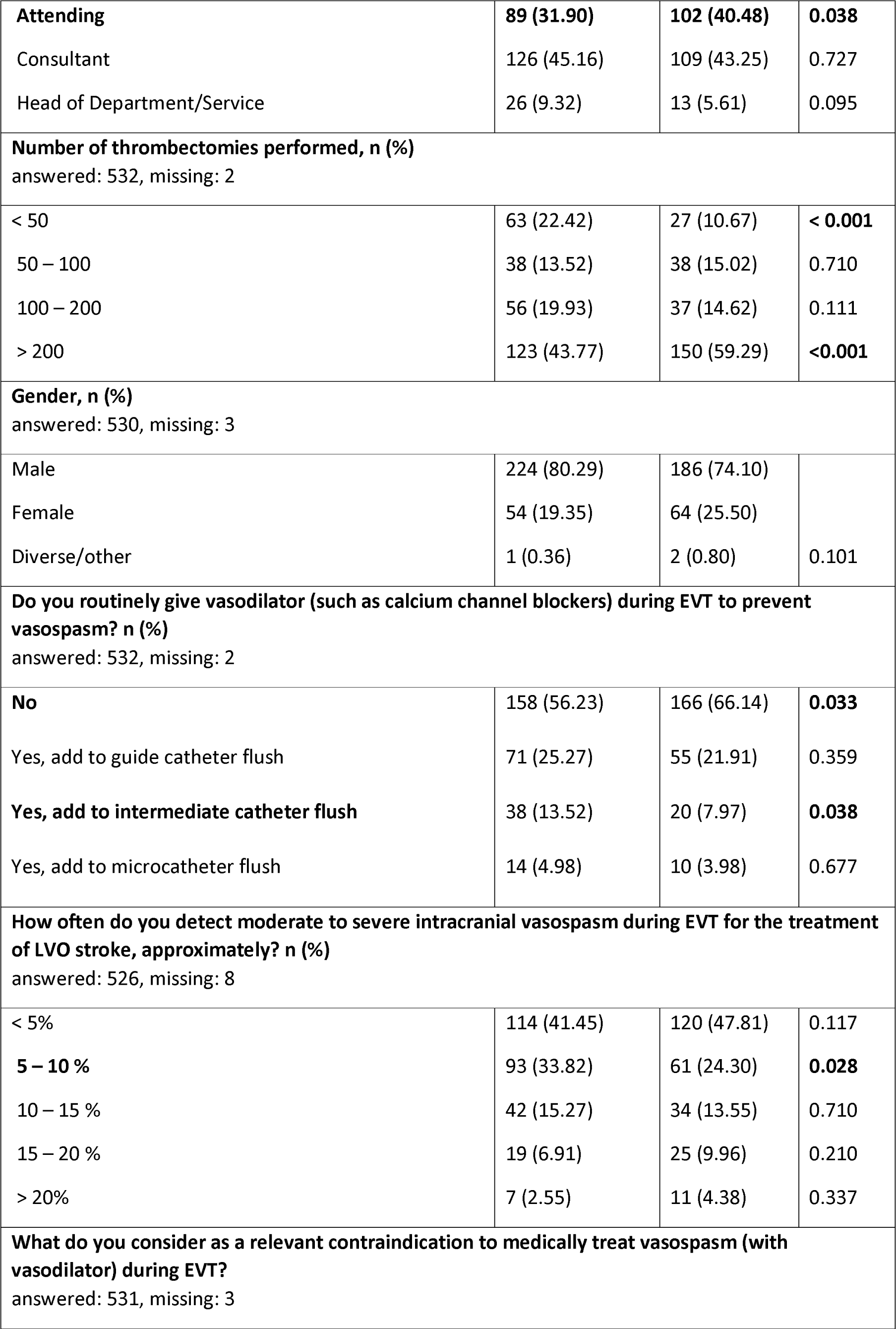
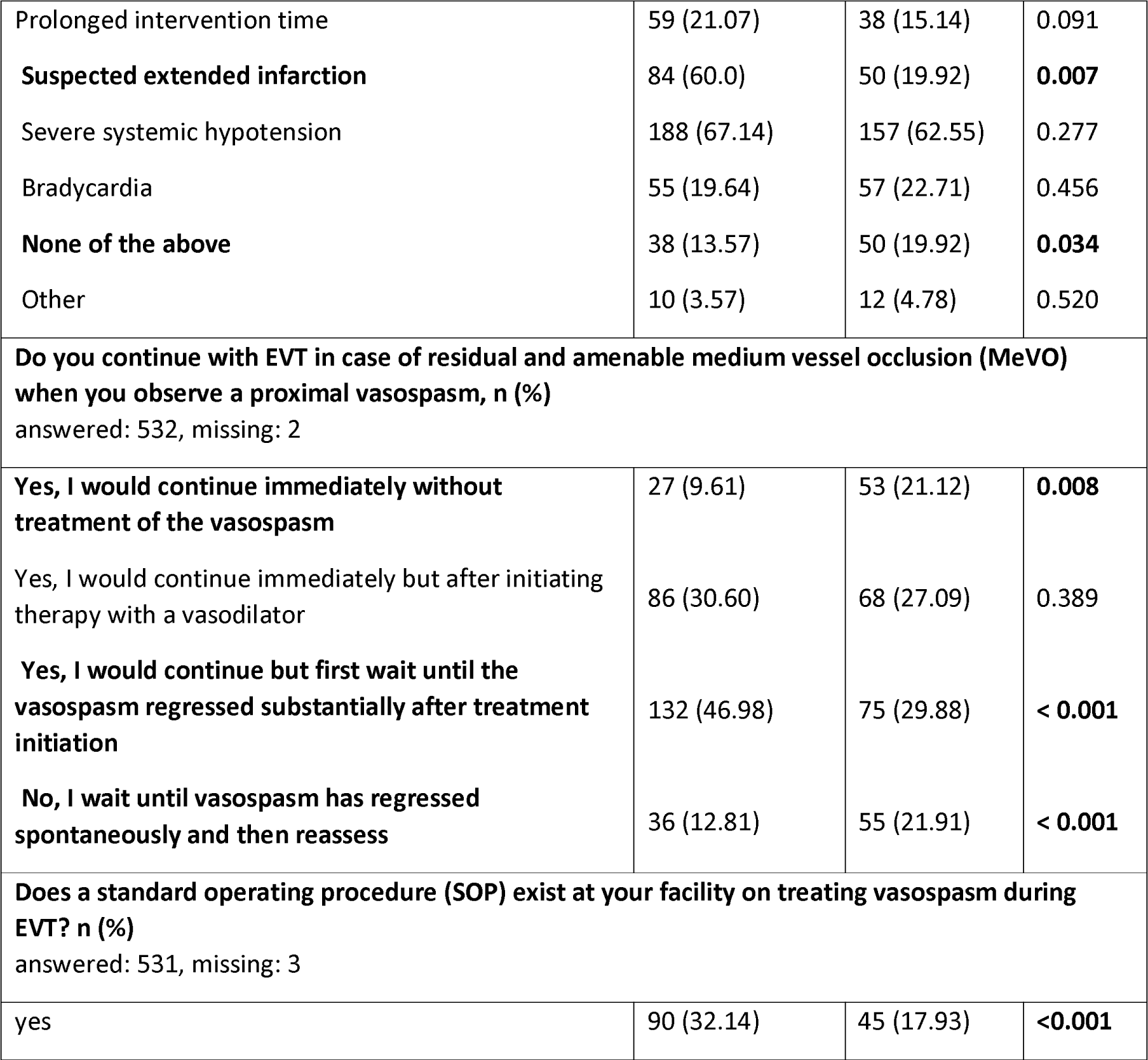
Group comparison according to the neuro-interventionalist opinion on potential influence of vasospasm regarding the stroke patient’s clinical outcome after EVT (Group 1: yes, vasospasm potentially influences the patient’s outcome, Group 2: no, vasospasm does not influence the patient’s outcome).

When participants were shown a large-vessel vasospasm in the left middle cerebral artery M1- and M2-segment (Figure 1A), Group 1 was more likely to treat it compared to Group 2 (75.3% vs. 56.0%, p < 0.001 Figure 1B). In the setting of a successful EVT and ensuing detection of large-vessel vasospasm, the majority of Group 1 would treat the vasospasm and control its regression with diagnostic angiographic imaging (Group 1: 74.3 % vs. 46.2 %, p < 0.001), while there was a high percentage in Group 2 who would withdraw the remaining EVT material without any further imaging or treatment (Group 1: 11.1% vs. Group 2: 36.3 %, p < 0.001) – Figure 1C. When shown a medium-vessel vasospasm in the left middle cerebral artery M3-segment (Figure 2A), Group 1 was more likely to treat it (27.1 % vs. 11.2 %; p < 0.001 – Figure 2B). This was also the case when shown an extracranial, hemodynamically not relevant vasospasm (Figure 3A) with a higher likelihood of treating the vasospasm in Group 1 compared to Group 2 (61.4% vs. 36.5 %, p < 0.001; Figure 3B).

**Figure 1.**
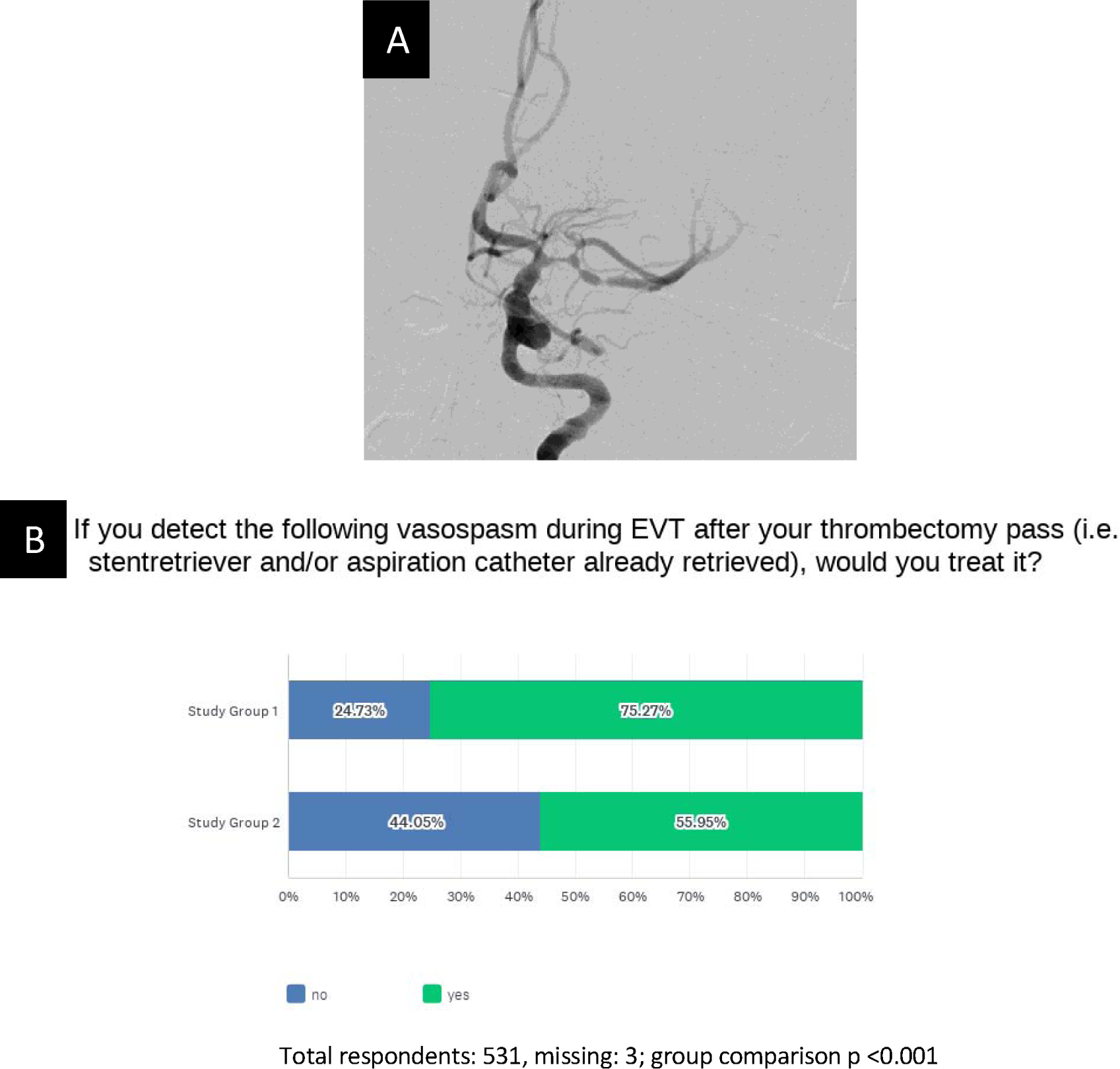
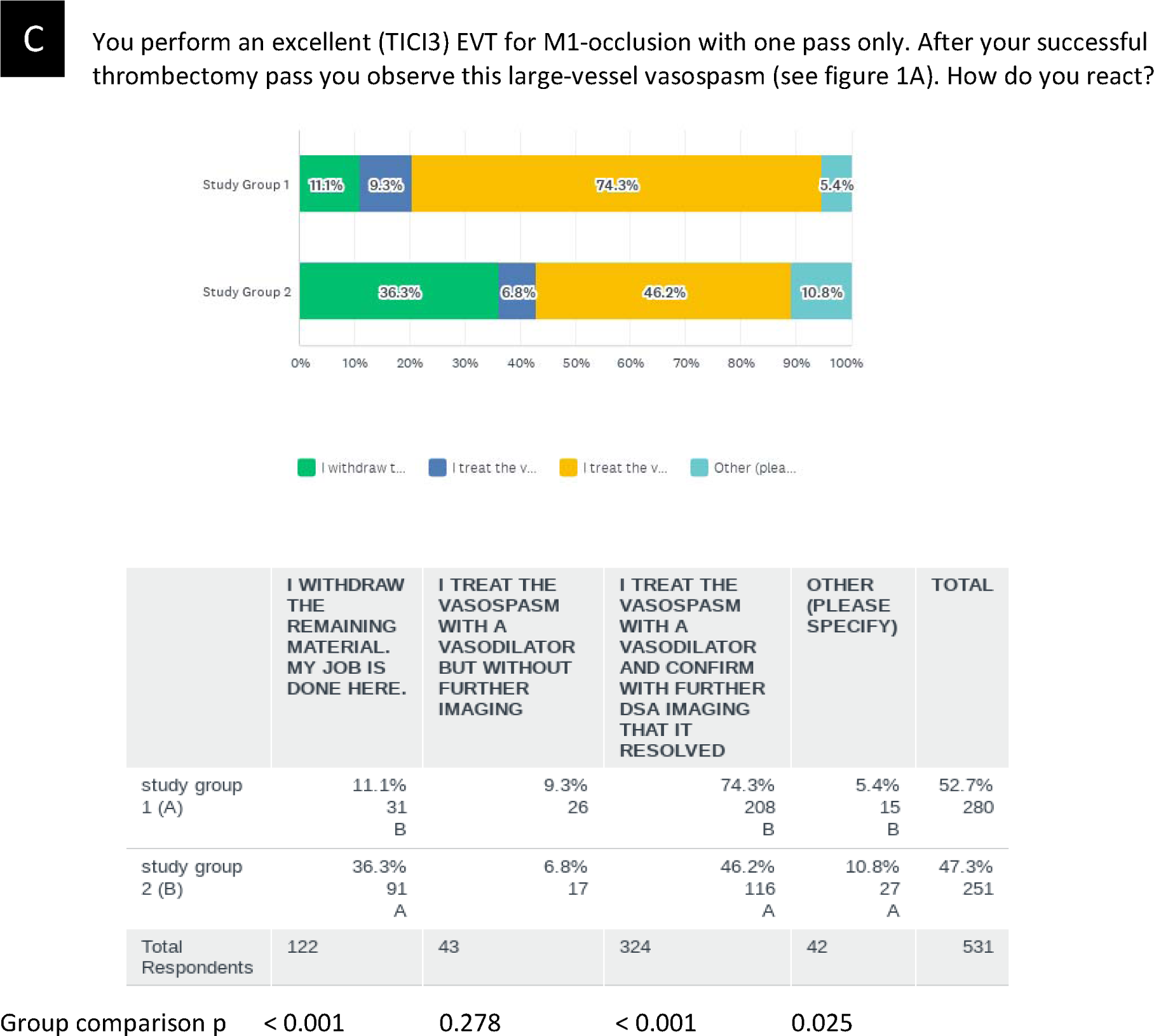
(A) Large-vessel vasospasm in the left middle cerebral artery M1- and M2-segments; (B) study group comparison on how to treat the depicted vasospasm and (C) how to continue an endovascular stroke treatment if this vasospasm is detected after successful EVT.

**Figure 2.**
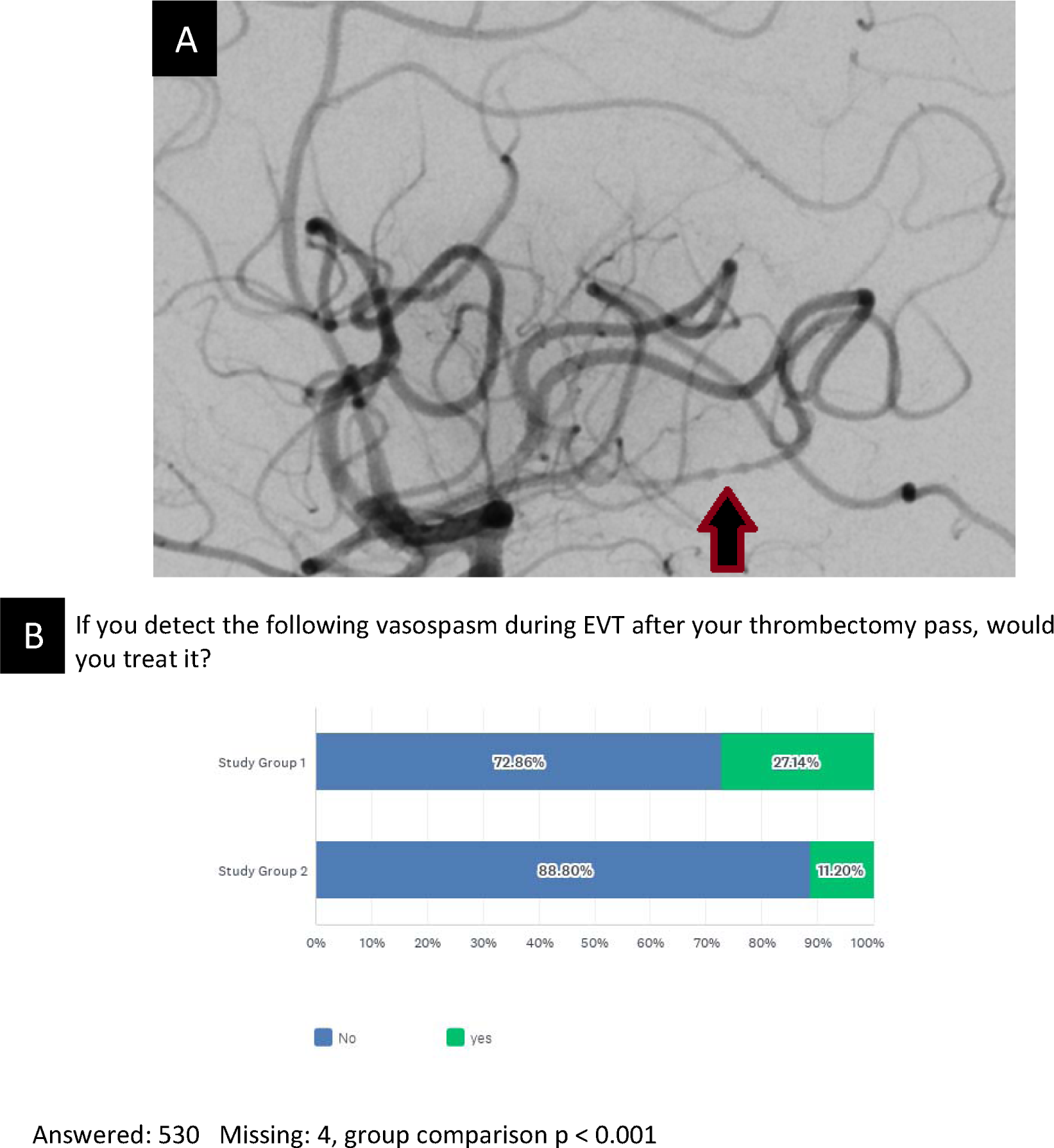
(A) Medium-vessel vasospasm in the left middle cerebral artery proximal M3-segment during endovascular stroke treatment (EVT); (B) study group comparison response on treatment of the vasospasm

**Figure 3.**
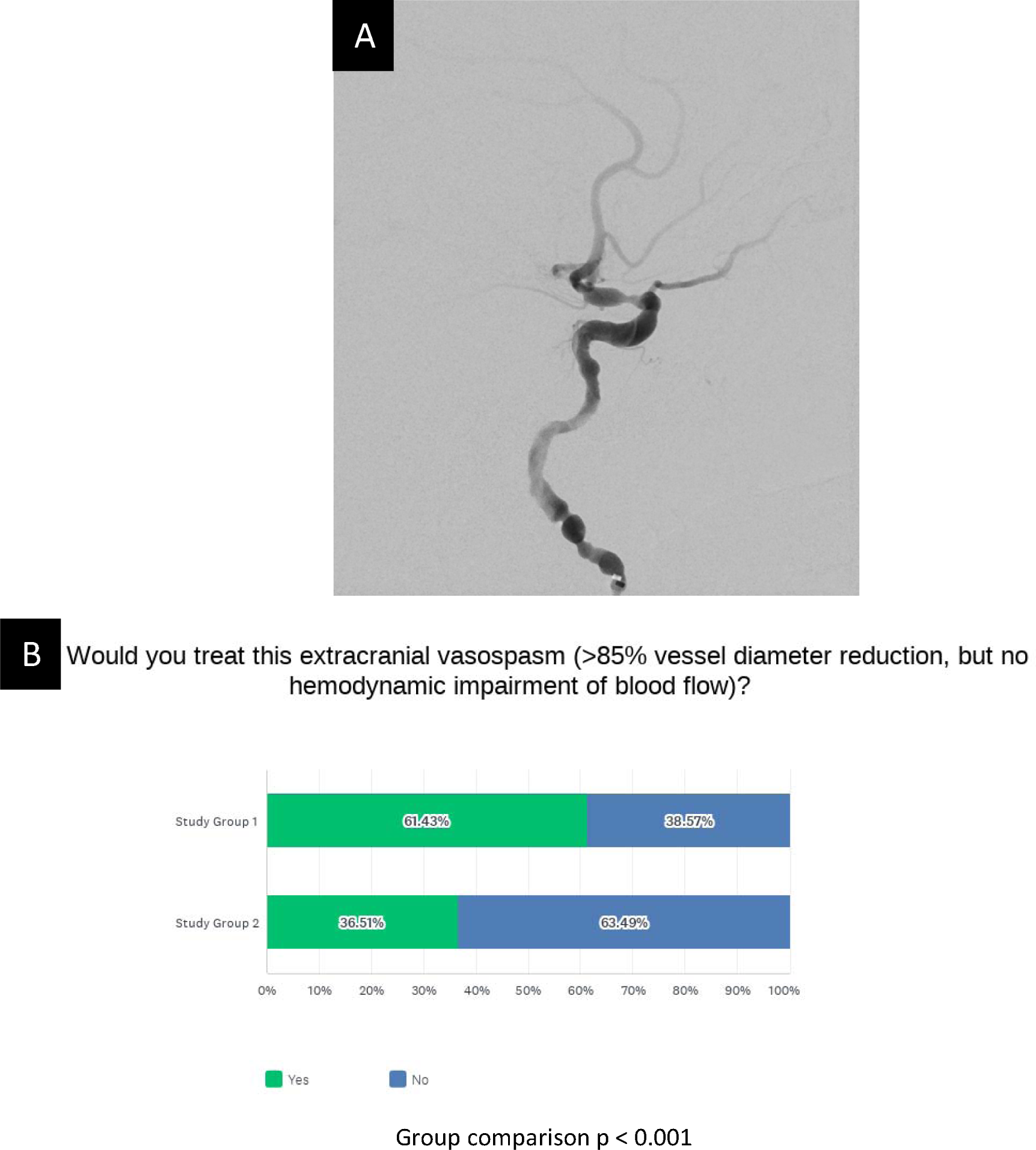
(A) Extracranial vasospasm of the left internal carotid artery C2-C6-segment (> 85% vessel diameter reduction, but no hemodynamic impairment of blood flow) during endovascular stroke treatment; (B) study group comparison of response on treatment of this vasospasm

## Discussion

This international online survey showed the differing opinion of neuro-interventionalists on the clinical relevance of vasospasm during EVT as well as heterogeneous treatment practice of vasospasm during EVT. While 52.6 % of respondents considered vasospasm as a complication after EVT that potentially influences the patient’s outcome (Group 1), 47.4 % of neuro-interventionalists did not share this view (Group 2).

The survey participants were highly experienced with over 200 EVTs performed in more than half of participants and over 10 years of neuro-interventional experience in 42%. There was no difference between groups regarding experience in the field or medical specialty (radiology, neuroradiology, surgery or neurology). The surveyed respondents in this analysis were from a diverse geographical background representing six continents and 56 countries. Female respondents were underrepresented with 22%, which reflects the current status of female underrepresentation in neurointervention and is therefore not to be seen as a study bias [14]. It has been shown in a Western-European cohort, that intracranial vasospasm during EVT for acute stroke was present in the range of 5-10% and was more likely to occur in younger patients [6]. The detection rate among respondents did not differ regarding study groups or geographical background of the NI. NIs practicing in Asia more often added vasodilators routinely to their flushes. Vasospasm was regarded less frequently as relevant complication by respondents practicing in North America while respondents practicing in Asia tended to regard vasospasm during EVT as relevant complication. It can be hypothesized that Asians could be more prone to the development of cerebral vasospasm, similar to what has been shown for coronary vasospasm [9], and are therefore more aware of vasospasms in cerebral vessels as a relevant complication and more willing to take countermeasures by routinely adding vasodilator to their flushes. To our knowledge potential population-based differences of the occurrence of cerebral vasospasms during EVT have not been studied yet.

Interestingly, the interventionalist’s opinion on the clinical relevance of vasospasm during EVT was associated with different treatment strategies. While we do not know if the presence of vasospasm during EVT can influence technical reperfusion success or the patient’s outcome, calcium channel blockers are known to reverse vasospasm in the majority of cases and some EVT patients experiencing vasospasm might show higher infarct volumes despite vasospasm being a transient phenomenon [12]. Future studies on this subject may be influenced by differing treatment strategies, as we learned from this study. Regarding a large-vessel vasospasm, the neurointerventionalists showed not only different opinions on treating the vasospasm, but more importantly, when and how to proceed with the EVT in case of a residual medium-vessel occlusion. While 49% of NI’s who considered vasospasm as a relevant complication would initiate therapy with intra-arterial vasodilators and continue with the EVT regardless of the vasospasm’s resolution, but only 29 % of NI’s who did not consider vasospasm as relevant complication initiate therapy before continuing. Time to recanalization is pivotal for patients with acute ischemic stroke and EVT, so that treatment initiation and related time delay might influence patient outcome, but also the risk of continuing EVT and probing in a spastic vessel is still unknown [15]. After a successful EVT with no remaining intracranial vessel occlusion but associated large-vessel vasospasm, 36% of interventionalists, who did not consider vasospasm during EVT a relevant complication, would withdraw their devices immediately without further imaging or therapy.

Further studies are necessary to evaluate the clinical significance of vasospasm during EVT, which will inform clinical practice.

## Limitations

This study is limited by the survey design and question selection. The questions in the survey pertained to vasospasm during acute stroke intervention. Given the time sensitive nature of acute stroke treatment, the results of this study may not apply to vasospasm treatment approaches in other neurovascular interventions such as endovascular aneurysm treatment. With categorical or ordered survey responses, the study allowed for quantitative analysis and a higher response rate due to a shorter time needed for responding to all survey questions. Free text responses could have offered a more detailed understanding of the interventionalist’s perspective and treatment practice but would have been difficult to quantify. An accurate response rate could not be ascertained because of overlapping members who are part of multiple organizations. The estimated response rate for survey participation was overall low at about 20% most likely due to broad survey distribution via email, which was deliberate to reach a broad and diverse audience. This relatively low response rate could have biased the study results. The survey’s completion rate however, was very high (99 %).

## Conclusion

The perceived clinical importance and management strategies of vasospasm during EVT varies considerably among neurointerventionalists. Physicians who believe that peri-interventional vasospasm does not influence clinical outcomes add vasodilators less often to catheter flushes, treat vasospasm less often when they occur and perform check angiograms less frequently. On the other hand, those who believe that vasospasm is detrimental to patient outcome often do inject vasodilators and perform angiographic controls to check for vasospasm resolution. More importantly, the experts’ opinion on the relevance of vasospasm also impacts the continuation of the thrombectomy per se. The heterogeneity in management strategies observed in our study reflects the lacking data on optimal vasospasm treatment strategies in the setting of EVT. This evidence gap should be addressed in future research.

## Supporting information

Supplement_CompleteSurvey

## Data Availability

Original data can be provided by the corresponding author upon reasonable request via eMail.

## Acknowledgements

We thank the German Society of Neuroradiology (Deutsche Gesellschaft für Neuroradiologie, DGNR, Bund Deutscher Neuroradiologen, BDNR) and the European Society of Minimally invasive Neurological Treatment (ESMINT) for making this survey visible by publishing it in their newsletter and sharing it with their email membership.

## References

1. Gascou, G., et al., Stent retrievers in acute ischemic stroke: complications and failures during the perioperative period. AJNR Am J Neuroradiol, 2014. 35(4): p. 734–40.

2. Nariai, Y., et al., Possible Contribution of the Aspiration Catheter in Preventing Post-stent Retriever Thrombectomy Subarachnoid Hemorrhage. Clin Neuroradiol, 2023. 33(2): p. 509–518.

3. Balami, J.S., et al., Complications of endovascular treatment for acute ischemic stroke: Prevention and management. Int J Stroke, 2018. 13(4): p. 348–361.

4. Pilgram-Pastor, S.M., et al., Stroke thrombectomy complication management. J Neurointerv Surg, 2021. 13(10): p. 912–917.

5. Behme, D., et al., Complications of mechanical thrombectomy for acute ischemic stroke-a retrospective single-center study of 176 consecutive cases. Neuroradiology, 2014. 56(6): p. 467–76.

6. Jesser, J., et al., Prediction and outcomes of cerebral vasospasm in ischemic stroke patients receiving anterior circulation endovascular stroke treatment. Eur Stroke J, 2023: p. 23969873231177766.

7. Saver, J.L., et al., Stent-retriever thrombectomy after intravenous t-PA vs. t-PA alone in stroke. N Engl J Med, 2015. 372(24): p. 2285–95.

8. Emprechtinger, R., B. Piso, and P.A. Ringleb, Thrombectomy for ischemic stroke: meta-analyses of recurrent strokes, vasospasms, and subarachnoid hemorrhages. J Neurol, 2017. 264(3): p. 432–436.

9. Woudstra, J., et al., Meta-analysis and systematic review of coronary vasospasm in ANOCA patients: Prevalence, clinical features and prognosis. Front Cardiovasc Med, 2023. 10: p. 1129159.

10. Pristipino, C., et al., Major racial differences in coronary constrictor response between japanese and caucasians with recent myocardial infarction. Circulation, 2000. 101(10): p. 1102–8.

11. Ducruet, A.F., et al., Genetic determinants of cerebral vasospasm, delayed cerebral ischemia, and outcome after aneurysmal subarachnoid hemorrhage. J Cereb Blood Flow Metab, 2010. 30(4): p. 676–88.

12. Jesser, J., et al., Effect of intra-arterial nimodipine on iatrogenic vasospasms during endovascular stroke treatment - angiographic resolution and infarct growth in follow-up imaging. BMC Neurol, 2023. 23(1): p. 5.

13. Sharma, A., et al., A Consensus-Based Checklist for Reporting of Survey Studies (CROSS). J Gen Intern Med, 2021. 36(10): p. 3179–3187.

14. Power, S., et al., Women in neurointervention, a gender gap? Results of a prospective online survey. Interv Neuroradiol, 2022. 28(3): p. 311–322.

15. He, A.H., et al., Every 15-min delay in recanalization by intra-arterial therapy in acute ischemic stroke increases risk of poor outcome. Int J Stroke, 2015. 10(7): p. 1062–7.

