## Supplement_CompleteSurvey for "Treatment practice of vasospasm during endovascular thrombectomy: an international survey"

**Supplemental Material**

*„ Therapy practice of vasospasm during endovascular stroke treatment (EVT)"*

An International anonymous online survey for neurointerventionalists

Vasospasm is a common complication during EVT of acute ischemic stroke, which occurs in 3% to 19% of EST cases. Although the description of vasospasm as a complication of endovascular stroke treatment traces back to as early as 2009, a current treatment paradigm for vasospasm does not exist. Therefore, we aim to investigate the international opinion of neurointerventionalists about the occurrence and management of vasospasm during EVT of acute ischemic stroke.

This anonymous international survey is not supported by any governmental or industrial funding. It is supported by the German Society of Neuroradiology (DGNR) and European Society of Minimally Invasive Neurological Therapy (ESMINT). We appreciate your help as neuro-interventionalists and community to help us gain an overview on world-wide standards regarding vasospasm therapy during EVT. The survey takes less than five minutes.

1.) In which country do you currently practice?
>> text field <<

2.) How many years of experience concerning endovascular stroke treatment do you have?
i) < 1 year
ii) 1-5 years
iii) 5-10 years
iv) > 10 years

3.) What is your specialty?
i) Interventional neuroradiologist
ii) Interventional neurologist
iii) Endovascular surgeon
iv) Interventional radiologist
v) other (please specify)

4.) What is your current position
i) fellow
ii) resident
iii) attending
iv) consultant
v) other (please specify)

5.) How many thrombectomies have you already performed?
i) < 10
ii) < 50
iii) 50-100
iv) 100-200

v) > 200

6.) What is your gender?
i) male
ii) female
iii) other/diverse

7.) Do you routinely give vasodilators (such as calcium channel blockers) during EVT to prevent vasospasm and if so, to which catheter flush do you add it?
i) Guide catheter (e.g. FlowGate, Mercy, Bobby, Walrus,…) flush
ii) Intermediate catheter (e.g. Sofia, Catalyst,…) flush
iii) Microcatheter (e.g. Rebar18, Headway17, Trak21,…) flush
iv) No, I do not routinely add calcium channel blockers (CCB) to any flush during EST

8.) If yes in Q7, which vasodilator do you normally use for the treatment of vasospasm during EVT?
i) Nimodipine
ii) Milrinone
iii) Verapamil
iv) Papaverin
v) other (please specify)

9.) If yes in Q7, how much milligram of which vasodilator do you add into how many millilitres flush?
>>> text field<<<

10.) How often do you approximately detect intracranial vasospasms during EST?
i) < 5%
ii) 5-10%
iii) 10-15%
iv) 15-20%
v) >20%

11.) If you detect the following intracranial vasospasm during EVT after your thrombectomy pass (i.e. stentretriever and/or aspiration catheter already retrieved), would you treat it?
i) – no
ii) – if yes, how much milligram of which vasodilator would you give over how many minutes? >>> text field <<<

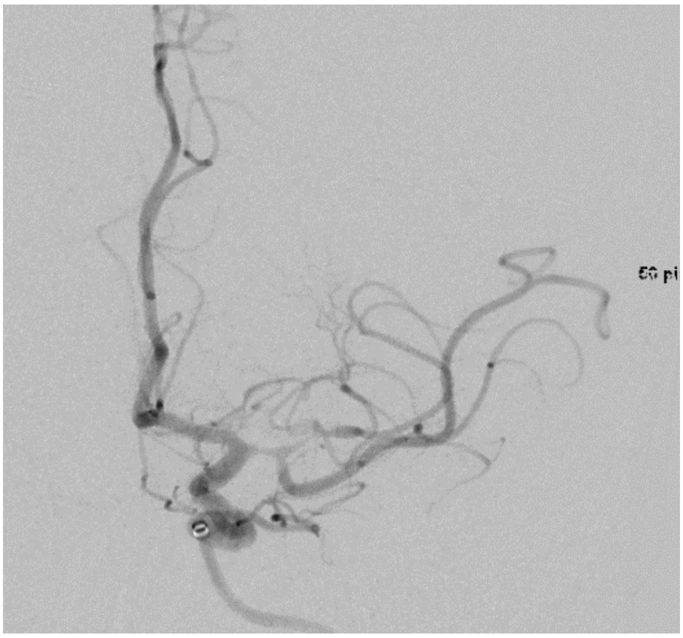

12.) Do you continue with EVT in case of residual and reachable medium vessel occlusion (MeVO), when you observe a proximal vasospasm such as shown in Q11?<

i) Yes, I would continue without treatment of the vasospasm
ii) Yes, I would continue immediately after I have initiated therapy with a vasodilator
iii) Yes, I would continue but first wait until the vasospasm regressed substantially after giving a vasodilator
iv) No, I wait until vasospasm has regressed spontaneously and then reassess

13.) You perform an excellent (TICI 3) EVT for M1-occlusion with one pass only! Congratulations! After your successful thrombectomy pass you observe a vasospasm such as shown in Q11. How do you react?

i) I withdraw the material. My job is done here.
ii) I treat the vasospasm with a vasodilator but without further imaging
iii) I treat the vasospasm with a vasodilator and confirm with further DSA imaging that it resolved
iv) other (please specify)

14.) If you detect the following intracranial vasospasm during EST, how much of which calcium channel blocker do you routinely give over how many minutes if at all?

i) no

ii) If yes, how much milligram of which vasodilator would you give over how many minutes?

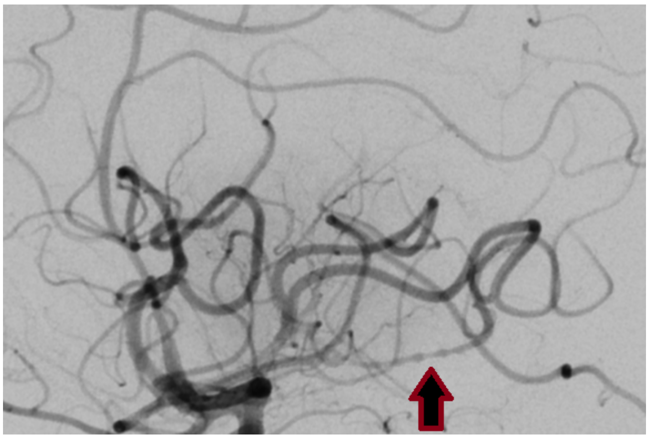

15.) do you consider intracranial vasospasm to be a complication that potentially influences the patient's outcome?
i) yes
ii) no

16.) Would you medically treat this extracranial vasospasms (> 85% vessel diameter reduction) ?

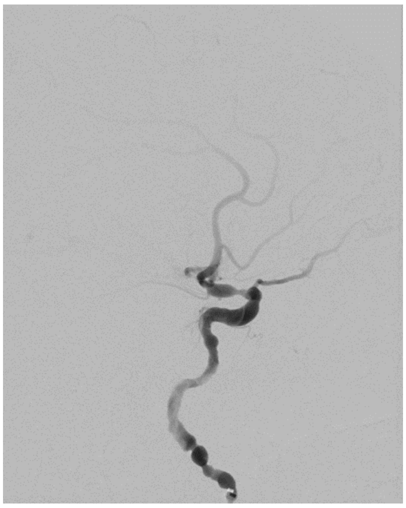

i) yes
ii) no

17.) What do you consider as relevant contraindication to medically treat vasospasms (with vasodilator) during EVT?
i) prolonged intervention time
ii) suspected extended infarction
iii) severe systemic hypotension
iv) bradycardia
v) other, please specify >> text field <<

18.) Does a standard operating procedure (SOP) exist at your facility on treating vasospasms during EVT?
i) yes
ii) no
